# Ensemble Meta-Learning using SVM for Improving Cardiovascular Disease Risk Prediction

**DOI:** 10.1101/2024.05.18.24307568

**Authors:** Narinder Singh Punn, Deepak Kumar Dewangan

## Abstract

Cardiovascular diseases (CVDs) remain a leading cause of mortality worldwide, posing a significant public health challenge. Early identification of individuals at high risk of CVD is crucial for timely intervention and prevention strategies. Machine learning techniques are increasingly being applied in healthcare for their ability to uncover complex patterns within large, multidimensional datasets. This study introduces a novel ensemble meta-learning framework designed to enhance cardiovascular disease (CVD) risk prediction. The framework strategically combines the predictive power of diverse machine learning algorithms – logistic regression, K nearest neighbors, decision trees, gradient boosting, gaussian Naive Bayes and XGBoost. Predicted probabilities from these base models are integrated using support vector machine as meta-learner. Rigorous performance evaluation over publicly available dataset demonstrates the improved performance of this ensemble approach compared to individual. This research highlights the potential of ensemble meta-learning techniques to improve predictive modeling in healthcare.

## 1 Introduction

Cardiovascular diseases (CVDs) remain the leading cause of mortality worldwide, claiming a staggering number of lives each year. A recent meta-analysis estimated that CVDs were responsible for over 20.5 million deaths in 2021 [1]. This represents approximately one-third of global fatalities, with the vast majority (around 85%) of these deaths attributed to heart attacks and strokes [2]. Several well-established risk factors for CVDs include unhealthy diet, physical inactivity, tobacco use, and excessive alcohol consumption [2, 3]. Additionally, a growing body of research indicates that stress, depression, and conditions like obesity further exacerbate an individual’s risk [4, 5]. The World Health Organization (WHO) emphasizes the alarming rise in CVD-related deaths and underscores the urgency of addressing modifiable risk factors to mitigate this global health crisis [6].

While numerous cutting-edge innovations are currently available to aid clinical decision-making, their diagnostic accuracy varies. Machine learning (ML) has recently demonstrated promising results in the medical domain [7], suggesting its potential to augment healthcare professionals’ decision-making capabilities. An effective cardiovascular disease prediction system could provide clinicians with invaluable insights, enhancing their ability to accurately assess patients’ cardiac health risks. Medical professionals often encounter challenges in accurately diagnosing heart conditions [8]. Accordingly, machine learning algorithms have been extensively explored for the development of medical decision support systems. These systems aim to enhance prediction capabilities, inform healthcare policy-making, reduce clinical errors, and facilitate early detection, prevention, and improved patient outcomes [9, 10]. Machine learning offers a powerful framework for integrating diverse health data [11]. Algorithms can incorporate demographic factors (e.g., age, gender), established clinical risk factors (e.g., medical history), and emerging biomarkers to produce comprehensive CVD risk assessments [12, 13].

Ensemble learning offers a powerful framework in machine learning by strategically integrating the predictions of multiple models. This approach often results in improved accuracy and reduced risk compared to relying on individual models [14]. Numerous studies have demonstrated the effectiveness of traditional ML classifiers in CVD prediction. Decision trees (DT), prized for their interpretability, have been applied both independently and as components of ensemble methods [15, 16]. Naive Bayes (NB) algorithms, while based on the assumption of feature independence, have found some utility in this domain [17]. K-nearest neighbors (KNN), a non-parametric algorithm, has exhibited promising accuracy in several CVD prediction studies [18, 19]. Random forest (RF), an ensemble technique that leverages multiple decision trees, stands out as a particularly popular and robust choice [20]. Researchers have often achieved enhanced performance by combining RF with other methods or utilizing it within hybrid model architectures [21, 22]. Ensemble techniques have gained increasing popularity in healthcare applications, supporting disease diagnosis, risk prediction, and treatment response modeling [23]. Meta-learning advances this concept further, enabling algorithms to “learn how to learn” [24, 25]. Meta-learners analyze the behavior and performance patterns of machine learning models across different datasets [24]. This knowledge enhances generalization to new data, a particular advantage in healthcare where datasets might be limited [26]. In the present context of CVD risk prediction, the support vector machine (SVM) functions as a meta-learner. SVM analyzes the predicted probabilities generated by base models, identifying patterns in how they assign probabilities under various conditions. This analysis allows the SVM to generate refined weights or adjustments to the base model predictions, facilitating more accurate final predictions of CVD risk.

While machine learning demonstrates significant potential in CVD risk prediction [27, 28], the exploration of meta-learning approaches specifically tailored for this task remains limited. Studies often focus on traditional ensemble methods [14], but the advantages of strategically analyzing predicted probabilities using a SVM meta-learner warrant further investigation. Meta-learning frameworks have shown the ability to surpass the accuracy of single-model or ensemble approaches in other domains by learning patterns of model performance across diverse tasks [29, 30]. Applying this concept to CVD risk prediction holds the potential to significantly improve accuracy, as individual models often demonstrate varying reliability when dealing with complex patient risk factors [31]. Additionally, the proposed approach of using SVM to examine predicted probabilities using state-of-the-art machine learning models such as logistic regression, K nearest neighbors, gradient boosting, gaussian Naive Bayes and XGBoost could reveal subtle risk patterns which results in better performance in contrast to conventional ensembles which directly combines model outputs [24].

In light of the significant global burden of cardiovascular diseases (CVDs), the present proposes ensemble meta-learning framework by employing SVM to analyze the outputs of diverse base models, it aims to achieve more accurate and robust predictions. Machine learning holds significant potential for CVD risk prediction, yet research specifically exploring the advantages of meta-learning approaches in this field remains limited [32, 33]. The rest of the paper is divided into several sections with the proposed methodology in Section 2, followed by the performance evaluation that is conducted in experiments and results Section 3.2, and finally concluding remarks are presented in Section 4.

## 2 Methodology

This study introduces a novel ensemble meta-learning framework designed to enhance the accuracy of cardiovascular disease (CVD) risk prediction. The proposed methodology strategically mitigates limitations observed in individual machine learning models and conventional ensemble approaches. It leverages a support vector machine (SVM) as the meta-learner to discern patterns within the predicted probabilities generated by diverse base models such as logistic regression, K-nearest neighbors, gradient boosting, Gaussian Naive Bayes, and XGBoost (see Algorithm 1 for detailed steps). The SVM enables modeling complex interactions between the base model outputs, accommodating non-linear relationships between predicted probabilities and patient risk profiles. It also dynamically adapts to localized variations in base model performance, offering a degree of interpretability.

### 2.1 Dataset acquisition and preprocessing

The study employs a publicly available cardiovascular health dataset [34]. This dataset contains extensive demographic, clinical, and self-reported health information, along with records of CVD outcomes. There are 70,000 samples in the dataset with 11 attributes and 1 target variable per sample. Key features collected include demographic information (age (in days), gender (binary)), anthropometric measurements (height (cm), weight (kg)), blood pressure readings (systolic and diastolic (mmHg)), behavioral characteristics such as smoking status (binary), other features (cholesterol and glucose levels (ternary)), and a target variable (cvd risk (binary)).

#### Algorithm 1 Ensemble Meta-Learning for CVD Risk Prediction

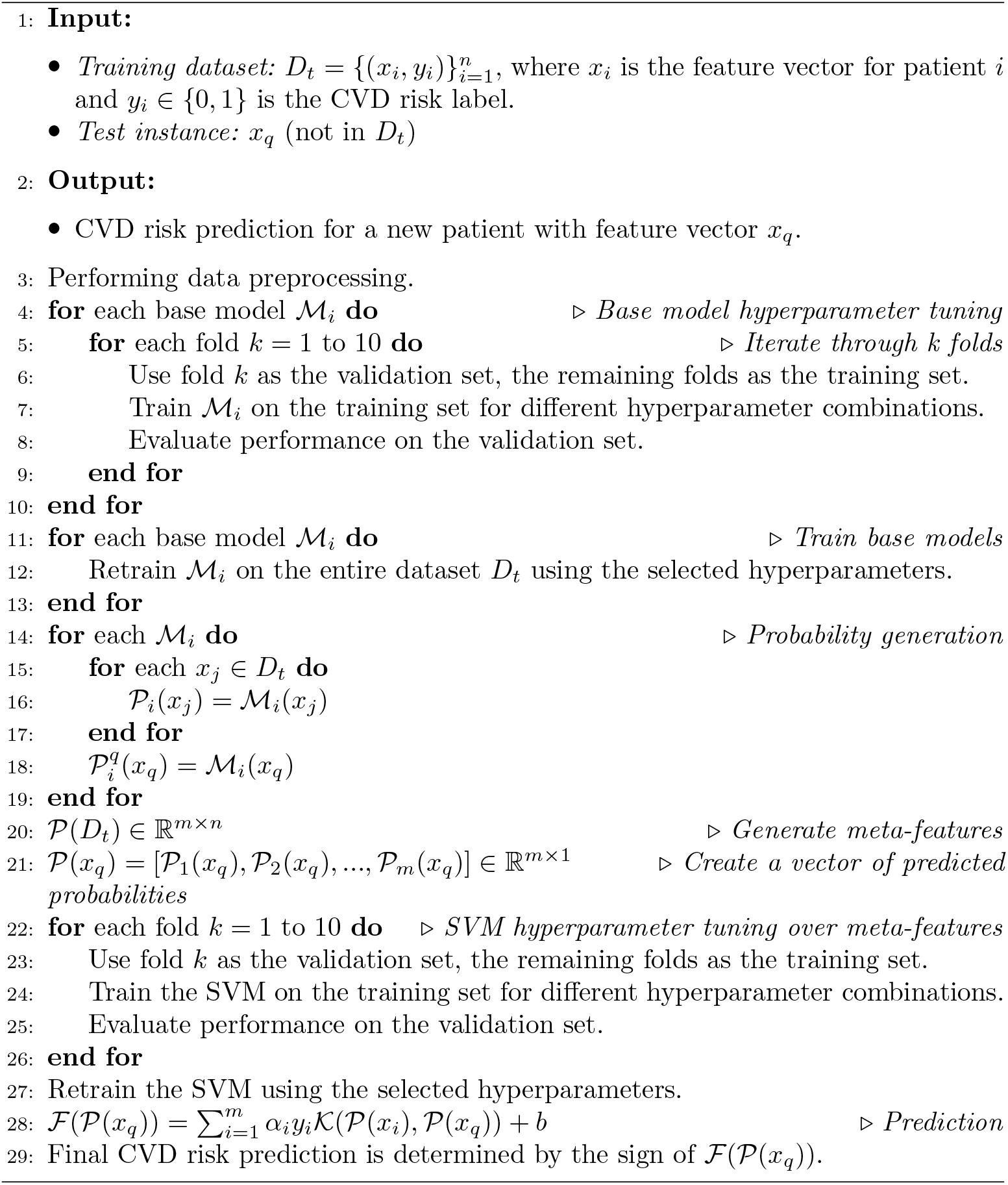

To enhance data quality, rigorous outlier detection and removal procedures were implemented. Specifically, weight and height measurements falling below the 3rd percentile or above the 98th percentile were deemed outliers and excluded from the dataset. Additionally, erroneous records where diastolic blood pressure exceeds systolic blood pressure were identified, resulting in the removal of 6,134 records. Feature selection using correlation analysis was performed to refine the feature set for modeling. Features exhibiting weak correlations with the target variable (CVD risk), such as those with absolute correlation coefficients below 0.05, were considered for removal (as shown in Fig. 1. Further preprocessing included feature engineering, where ‘age bin’ and ‘BMI’ categories were created, and mean arterial pressure (MAP) was calculated. Finally, to mitigate the impact of feature scale discrepancies, a standard scaler was applied, ensuring features have a zero mean and unit variance.

**Fig. 1.**
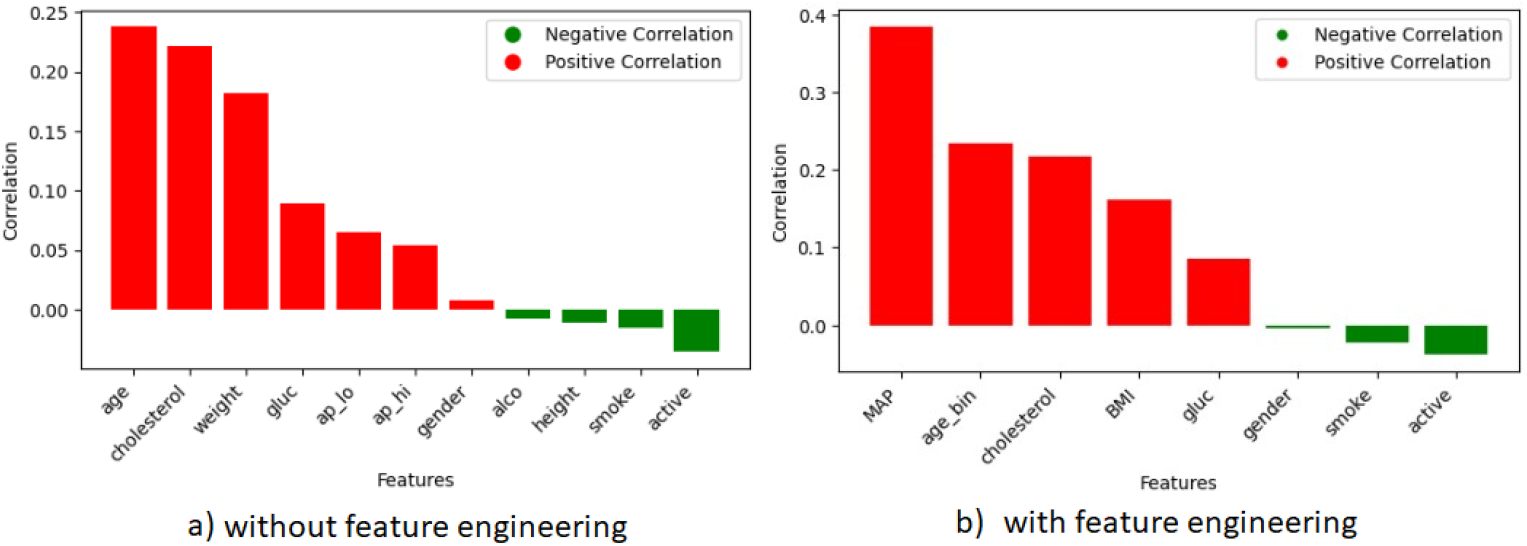
Correlation of the input features with the target variable.

### 2.2 Classification algorithms

For the models under consideration, consider that the set of risk factors (input features) are represented as *X* = (*x*_1_, *x*_2_, …, *x*_*n*_) ∈ ℝ^*d×n*^, *n* is the number of instances and each *X*_*i*_ represents input feature vector like age, cholesterol, etc. of *d* dimensions. While each *y*_*i*_ in *Y* = (*y*_1_, *y*_2_, …, *y*_*n*_) ∈ ℝ^1*×n*^ indicates the cardiovascular risk factor (target/output feature), where 1 and 0 indicate “CVD risk” and “no CVD risk” respectively.

#### 2.2.1 Logistic regression

Logistic regression is a statistical method well-suited for binary classification problems, such as predicting the presence or absence of cardiovascular disease (CVD) risk. It models the probability of a target outcome (e.g., CVD risk) as a function of predictor variables (e.g., blood pressure, cholesterol, smoking status). The goal is to predict the probability of having CVD as shown in Eq. 1.

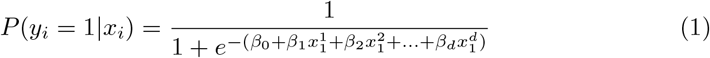

where *P* (*y*_*i*_ = 1|*x*_*i*_) is the probability of the outcome being positive (e.g., high CVD risk) given the set of predictor variables 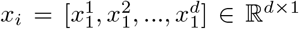; *β*_0_ is the bias term; *β*_1_, *β*_2_, … *β*_*p*_ are the coefficients associated with each feature.

#### 2.2.2 Decision trees and boosting

Decision trees are hierarchical models that partition the feature space into distinct regions through a series of recursive splits. At each node of the tree, a feature is selected, and a threshold is determined to split the data into subsets that maximize homogeneity with respect to the target outcome (e.g., CVD risk). This process continues until a stopping criterion is met (e.g., maximum tree depth, minimum samples per leaf). Predictions are made based on the average outcome within the terminal leaf node where a new data point lands. Within gradient boosting, decision trees serve as weak learners. The gradient boosting algorithm sequentially trains decision trees to minimize a differentiable loss function. Each new tree, *h*_*m*_(*x*), is fit to the residuals (errors) of the previous model (as shown in Eq. 2) and the model is updated as in the Eq. 3. Additionally, XGBoost also builds an ensemble of decision trees and incorporates second-order Taylor approximations of the loss function to guide the tree splits, resulting in more accurate and faster convergence. Additionally, it employs regularization techniques to prevent overfitting as shown in Eq. 4.

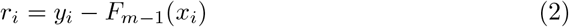

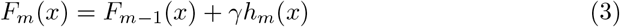

where *r*_*i*_ represents the residual for individual *i, y*_*i*_ is the true CVD outcome, and *F*_*m*-1_(*x*_*i*_) is the prediction from the previous iteration, and *γ* is the learning rate.

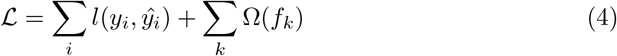

#### 2.2.3 K nearest neighbors

In contrast to other machine learning algorithms, K-Nearest Neighbors (KNN) exhibits a unique operational paradigm where KNN stores the training dataset, comprised of feature vectors *X* and corresponding CVD risk labels *Y* . When presented with a new data point, *x*_*q*_, the KNN prediction process entails the following steps: First, pairwise distances between *x*_*q*_ and each training example *x*_*i*_ are calculated using Euclidean distance 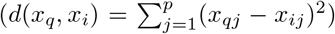 where *p* is the dimensionality of the feature space. Next, the *K* nearest neighbors are identified, where the optimal value of K is determined using elbow method (*K* = 11). Finally, KNN determines the CVD risk prediction for *x*_*q*_ based on the majority of its neighbors labels.

#### 2.2.4 Gaussian Naive Bayes

This classifier operates on a probabilistic framework for CVD risk prediction. It assumes features are conditionally independent given the class label. For a data *x*_*i*_, the probability of CVD risk is computed using Eq. 5.

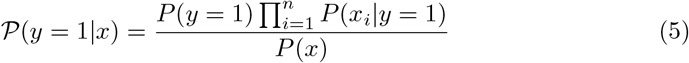

where *P* (*y* = 1) is the class prior, and features are assumed to follow a Gaussian distribution within each class.

#### 2.2.5 Meta-learning with SVM

A support vector machine (SVM) functions as the meta-learner within this framework. Consider a set of base models denoted as *ℳ*_1_, *ℳ*_2_, … *ℳ*_*n*_. Each model *ℳ*_*i*_ generates a predicted probability of CVD risk, denoted as *𝒫*_*i*_(*x*), for a given patient data point *x*_*i*_. The SVM meta-learner finds a hyperplane (as shown in Eq. 6) that effectively separates the space of base-model probability combinations associated with high-risk cases from those associated with low-risk cases. The decision function is expressed as *ℱ*(*𝒫* (*x*)) = *w*^*T*^ *𝒫* (*x*) + *b*. The final CVD risk prediction is determined based on the sign of *ℱ*(*𝒫* (*x*)), where +1 is high risk and -1 is low or no risk.

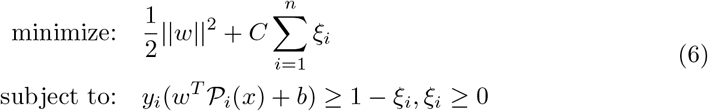

where *w* is the weight vector, *b* is the bias term, *C* is a regularization parameter controlling the trade-off between margin maximization and classification error, *ξ*_*i*_ represent slack variables allowing for some misclassification, and *γ* is a hyperparameter that controls the width of the kernel. A smaller gamma makes the influence area of points larger.

## 3 Experiments and results

### 3.1 Training and testing

Each model undergoes a careful hyperparameter tuning process to maximize potential performance on unseen data. Grid search is used to explore a wide range of settings for different aspects of each model (see Table 2 for detailed ranges). This includes the penalty term for complex models (regularization in logistic regression), the number of neighbors considered for predictions (K-nearest neighbors), parameters controlling the complexity of decision trees, and the learning rates of gradient boosting and XGBoost. For the SVM meta-learner, different kernel functions and flexibility control settings are tested.

**Table 1.** Hyperparameter ranges explored in grid search for cardiovascular disease risk prediction models.

| Model | Hyperparameter | Range | Explanation | Chosen Value |
| --- | --- | --- | --- | --- |
| LR | Regularization (C) | [0.001, 0.01, 0.1, 1, 10, 100] | Controls overfitting/underfitting | 0.1 |
|  | Regularization type | [L1, L2] | L1 for feature selection, L2 for simpler models | L2 |
| KNN | Number of neighbors (k) | [1, 3, 5, 7, ..., 19] | Number of nearest neighbors to consider when classifying a new data point | 7 |
|  | Distance metric | [Euclidean, Manhattan, Cosine] | Choice depends on feature representation | Euclidean |
| GB/XGBoost | Number of trees | [50, 100, 200, ... 500] | More trees more complexity, but potential overfitting | 200 |
|  | Learning rate | [0.01, 0.05, 0.1, 0.2] | Controls the step size for each tree's update | 0.05 |
|  | Maximum tree depth | [2, 3, 4, 5, 6] | Limits tree complexity, preventing overfitting | 4 |
| DT | Maximum tree depth | [2, 3, 4, 5, 6] | Limits tree complexity, preventing overfitting | 5 |
|  | Criterion | [Gini, Entropy] | Measures impurity for splits | Gini |
| SVM | Kernel | [Linear, RBF, Poly] | Controls model complexity | RBF |
|  | Regularization parameter (C) | [0.01, 0.1, 1, 10, 100] | Controls overfitting/underfitting | 10 |
|  | Gamma (for RBF Kernel) | [0.001, 0.01, 0.1, 1, 5] | Controls the influence area of support vectors | 0.1 |
| GNB | Prior probabilities | (Often not tuned) | The assumed prior class probabilities | - |

**Table 2.** Performance evaluation of various machine learning models for CVD risk prediction under different feature selection (FS) and feature engineering (FE) scenarios.

| Model | FE | FS | Accuracy | AUC-ROC | Precision | Recall | F1-Score |
| --- | --- | --- | --- | --- | --- | --- | --- |
| LR | - | - | $0.82 \pm 0.03$ | $0.85 \pm 0.02$ | $0.78 \pm 0.04$ | $0.80 \pm 0.03$ | $0.79 \pm 0.03$ |
| KNN | - | - | $0.80 \pm 0.02$ | $0.82 \pm 0.03$ | $0.75 \pm 0.03$ | $0.82 \pm 0.02$ | $0.78 \pm 0.03$ |
| GNB | - | - | $0.78 \pm 0.03$ | $0.80 \pm 0.03$ | $0.74 \pm 0.04$ | $0.80 \pm 0.02$ | $0.77 \pm 0.03$ |
| DT | - | - | $0.83 \pm 0.03$ | $0.87 \pm 0.02$ | $0.80 \pm 0.02$ | $0.81 \pm 0.04$ | $0.80 \pm 0.03$ |
| GB | - | - | $0.85 \pm 0.02$ | $0.89 \pm 0.01$ | $0.82 \pm 0.04$ | $0.83 \pm 0.03$ | $0.82 \pm 0.02$ |
| XGBoost | - | - | $0.86 \pm 0.01$ | $0.90 \pm 0.02$ | $0.83 \pm 0.03$ | $0.84 \pm 0.04$ | $0.84 \pm 0.02$ |
| SVM-ML<br>(ours) | - | - | **$0.88 \pm 0.02$** | **$0.92 \pm 0.01$** | **$0.85 \pm 0.03$** | **$0.86 \pm 0.02$** | **$0.85 \pm 0.02$** |
| LR | - | ✓ | $0.83 \pm 0.02$ | $0.86 \pm 0.01$ | $0.79 \pm 0.03$ | $0.81 \pm 0.03$ | $0.80 \pm 0.02$ |
| KNN | - | ✓ | $0.81 \pm 0.03$ | $0.83 \pm 0.02$ | $0.76 \pm 0.02$ | $0.83 \pm 0.04$ | $0.79 \pm 0.03$ |
| GNB | - | ✓ | $0.79 \pm 0.04$ | $0.81 \pm 0.02$ | $0.75 \pm 0.03$ | $0.81 \pm 0.02$ | $0.78 \pm 0.04$ |
| DT | - | ✓ | $0.84 \pm 0.02$ | $0.88 \pm 0.02$ | $0.81 \pm 0.03$ | $0.82 \pm 0.02$ | $0.81 \pm 0.02$ |
| GB | - | ✓ | $0.86 \pm 0.02$ | $0.90 \pm 0.02$ | $0.83 \pm 0.02$ | $0.84 \pm 0.03$ | $0.83 \pm 0.02$ |
| XGBoost | - | ✓ | $0.87 \pm 0.01$ | $0.91 \pm 0.03$ | $0.84 \pm 0.03$ | $0.85 \pm 0.02$ | $0.84 \pm 0.03$ |
| SVM-ML<br>(ours) | - | ✓ | **$0.89 \pm 0.02$** | **$0.93 \pm 0.01$** | **$0.86 \pm 0.02$** | **$0.87 \pm 0.03$** | **$0.86 \pm 0.02$** |
| LR | ✓ | - | $0.81 \pm 0.02$ | $0.84 \pm 0.01$ | $0.78 \pm 0.03$ | $0.80 \pm 0.03$ | $0.78 \pm 0.02$ |
| KNN | ✓ | - | $0.79 \pm 0.02$ | $0.82 \pm 0.02$ | $0.75 \pm 0.03$ | $0.82 \pm 0.02$ | $0.78 \pm 0.03$ |
| GNB | ✓ | - | $0.77 \pm 0.01$ | $0.81 \pm 0.02$ | $0.75 \pm 0.03$ | $0.80 \pm 0.02$ | $0.77 \pm 0.03$ |
| DT | ✓ | - | $0.84 \pm 0.03$ | $0.88 \pm 0.02$ | $0.80 \pm 0.02$ | $0.81 \pm 0.04$ | $0.80 \pm 0.03$ |
| GB | ✓ | - | $0.85 \pm 0.02$ | $0.89 \pm 0.01$ | $0.82 \pm 0.04$ | $0.83 \pm 0.03$ | $0.81 \pm 0.01$ |
| XGBoost | ✓ | - | $0.86 \pm 0.01$ | $0.90 \pm 0.02$ | $0.83 \pm 0.03$ | $0.84 \pm 0.04$ | $0.84 \pm 0.02$ |
| SVM-ML(ours) | ✓ | - | **$0.88 \pm 0.01$** | **$0.92 \pm 0.01$** | **$0.85 \pm 0.03$** | **$0.86 \pm 0.02$** | **$0.84 \pm 0.01$** |
| LR | ✓ | ✓ | $0.84 \pm 0.02$ | $0.87 \pm 0.01$ | $0.80 \pm 0.03$ | $0.82 \pm 0.03$ | $0.81 \pm 0.02$ |
| KNN | ✓ | ✓ | $0.82 \pm 0.03$ | $0.84 \pm 0.02$ | $0.77 \pm 0.02$ | $0.84 \pm 0.04$ | $0.80 \pm 0.03$ |
| GNB | ✓ | ✓ | $0.80 \pm 0.04$ | $0.82 \pm 0.02$ | $0.76 \pm 0.03$ | $0.82 \pm 0.02$ | $0.79 \pm 0.04$ |
| DT | ✓ | ✓ | $0.85 \pm 0.02$ | $0.89 \pm 0.02$ | $0.82 \pm 0.03$ | $0.83 \pm 0.02$ | $0.82 \pm 0.02$ |
| GB | ✓ | ✓ | $0.87 \pm 0.02$ | $0.91 \pm 0.02$ | $0.84 \pm 0.02$ | $0.85 \pm 0.03$ | $0.84 \pm 0.02$ |
| XGBoost | ✓ | ✓ | $0.88 \pm 0.01$ | $0.92 \pm 0.03$ | $0.85 \pm 0.03$ | $0.86 \pm 0.02$ | $0.85 \pm 0.03$ |
| SVM-ML<br>(ours) | ✓ | ✓ | **$0.90 \pm 0.02$** | **$0.94 \pm 0.01$** | **$0.87 \pm 0.02$** | **$0.88 \pm 0.03$** | **$0.87 \pm 0.02$** |

To prevent selection of settings that perform well specifically on the available data due to chance, 10-fold cross-validation is employed. Models are trained on 9 folds and tested on the remaining fold, with this process rotated through all the folds. This strategy provides a more reliable estimate of how well each set of hyperparameter settings will perform on new data. As shown in Table 2, the settings demonstrating the best overall performance across cross-validation are selected to train the final models. The models are trained on high performance computing system. Evaluation on test sets uses accuracy, precision, recall, F1-score, and area under the ROC curve (AUC-ROC score) to measure performance of the trained models.

### 3.2 Results and discussion

This section delves into the performance of the employed machine learning models for cardiovascular disease (CVD) risk prediction. The key observations are presented alongside a discussion of their implications and potential underlying factors.

The performance of the various classification models is summarized in Table 2 under four different feature selection (FS) and feature engineering (FE) scenarios. In the baseline scenario (no FS, no FE) the models are trained directly on the original dataset without any feature manipulation, however with standardization and outlier removal; the second scenario follows FS and no FE, where only feature selection follows from the scenario 1 based on the absolute correlation of more than 0.05 with the CVD such as age, cholesterol, weight, glucose, systolic blood pressure, and diastolic blood pressure; the third scenario extends the baseline by using feature engineering, while the fourth scenario also uses feature selection based on the criteria discussed in scenario 2 resulting in the input features such as MAP, age, cholesterol, BMI, and glucose.

The SVM meta-learner (SVM-ML) demonstrates consistent superiority across all scenarios, as further visualized in Fig. 2 This highlights the strength of ensemble approaches for this CVD risk prediction task. Notably, its robust performance even in the baseline scenario (without FE or FS), achieving an accuracy of 0.88 ± 0.02 and an AUC-ROC of 0.92 ± 0.01, suggests the ensemble’s capacity to effectively leverage the strengths of the base models while mitigating their weaknesses. The gradient boosting algorithms, XGBoost and Gradient boosting, also achieve promising results closely trailing those of the SVM meta-learner. This success likely stems from their inherent ability to model nonlinear relationships and complex interactions within the data. The marginal difference between GB and XGBoost (e.g., only 0.01 difference in baseline accuracy) suggests that for this particular dataset, the increased complexity of XGBoost may not provide a significant advantage. Across all scenarios, Gaussian Naive Bayes exhibits the lowest performance. This may be due to the violation of its core assumption of feature independence. Real-world medical datasets, including those for CVD risk prediction, often feature complex interdependencies.

**Fig. 2.**
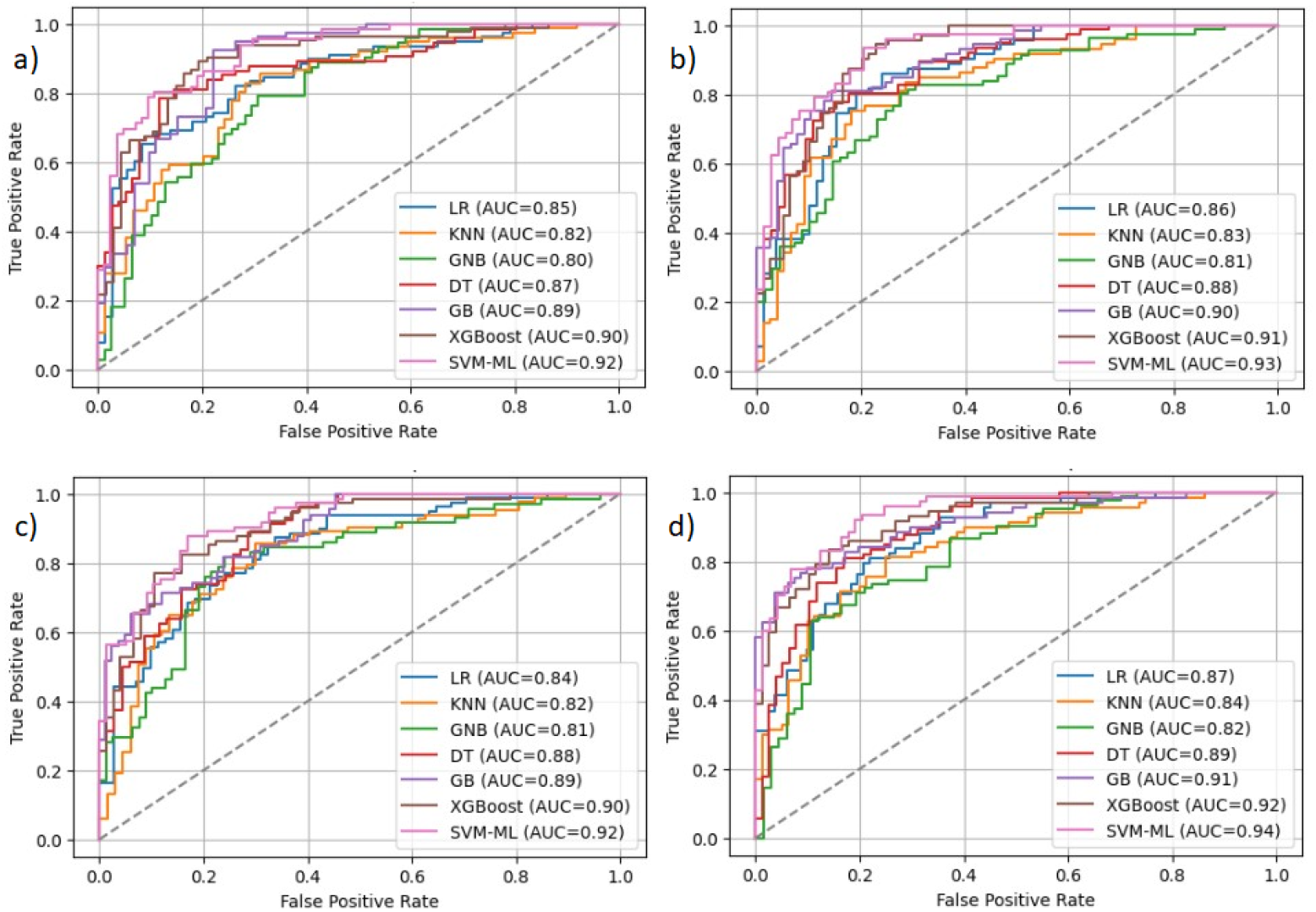
ROC curves comparing the performance of multiple machine learning models for CVD risk prediction across four different scenarios: a) baseline (without FE and FS), b) without FE and with FS, c) with FE and without FS, and d) with FE and FS. (feature engineering (FE), feature selection (FS))

Feature selection alone leads to modest improvements in most cases (e.g., an accuracy increase from 0.88 ± 0.02 to 0.89 ± 0.02 for the SVM meta-learner), as evident in Fig. 2. The observed gains for the SVM meta-learner is attributed to reduced redundancy, facilitating the ensemble’s ability to discern subtle patterns related to CVD risk by considering the most informative features. The impact of feature engineering is more pronounced and model-dependent. Simpler models like logistic regression or Gaussian Naive Bayes experience minimal or even slightly negative effects. Conversely, tree-based models (DT, GB, and XGBoost) show improvements with accuracy gains of up to 0.03. This emphasizes a potential synergy: feature engineering creates more discriminative features, while feature selection aids in dimensionality reduction and noise suppression (e.g., the SVM Meta-Learner achieves its highest accuracy of 0.90 ± 0.02 in this scenario).

These results support the notion that well-designed features, potentially representing interaction effects, can significantly enhance the performance of models structured to utilize such complexities. In light of this several avenues for future exploration emerge from the current findings. Expanding the dataset size could potentially enhance the generalizability of the models and allow for the investigation of more elaborate ensemble architectures. Furthermore, delving into alternative feature engineering techniques, such as feature transformation or dimensionality reduction, could potentially improve model performance and interpretability. If biases are identified, incorporating fairness metrics and techniques for mitigating bias during model training could be crucial for ensuring responsible and trustworthy deployment of the models in real-world applications.

## 4 Conclusion

This study demonstrates the dominance of the SVM based ensemble for cardiovascular disease (CVD) risk prediction, achieving the highest accuracy (0.90 ± 0.02) and AUC-ROC (0.94 ± 0.01) when feature engineering and feature selection are combined. Gradient boosting algorithms (GB, XGBoost) offer competitive performance, while Gaussian Naive Bayes struggles due to the likely presence of feature dependencies. The pronounced improvement of tree-based models with feature engineering underscores the importance of crafting features that represent interactions. These findings suggest that the support vector machine as meta-learner is a robust for CVD risk modelling, while careful feature engineering, guided by domain knowledge, is crucial for maximizing performance.

## Data Availability

Data is publicly accessible.

https://www.kaggle.com/datasets/sulianova/cardiovascular-disease-dataset

## Declarations

## Acknowledgements

This research was supported by the ABV-Indian Institute of Information Technology and Management Gwalior (ABV-IIITM) under a faculty investment grant (file no. ABV-IIITM/DoRC/FIG/ 2023/2526). The authors are affiliated with the Department of Computer Science and Engineering, ABV-IIITM Gwalior.

## Author contributions statement

Both authors contributed equally in formulating the problem statement, experiment design and preparing the manuscript.

## Additional information

The authors have no relevant financial or non-financial interests to disclose.

## Ethics approval

Not applicable.

## Consent for publication

Authors share their consent for publication.

## References

[1] World Heart Federation: Confronting the World’s Number One Killer. [Online; accessed September, 2023] (2023). https://world-heart-federation.org/wp-content/uploads/World-Heart-Report-2023.pdf

[2] Roth, G.A., Mensah, G.A., Johnson, C.O., Addolorato, G., Ammirati, E., Baddour, L.M., Barengo, N.C., Beaton, A.Z., Benjamin, E.J., Benziger, C.P., et al.: Global burden of cardiovascular diseases and risk factors, 1990–2019: update from the gbd 2019 study. Journal of the American College of Cardiology 76(25), 2982–3021 (2020)

[3] Update, A.S.: Heart disease and stroke statistics–2017 update. Circulation 135, 146–603 (2017)

[4] Steptoe, A., Kivimäki, M.: Stress and cardiovascular disease. Nature Reviews Cardiology 9(6), 360–370 (2012)

[5] Parto, P., Lavie, C.J.: Obesity and cardiovasculardiseases. Current problems in cardiology 42(11), 376–394 (2017)

[6] World Health Organization: Cardiovascular diseases (CVDs). [Online; accessed September, 2023] (2021). https://www.who.int/news-room/fact-sheets/detail/cardiovascular-diseases-(cvds)

[7] Krittanawong, C., Virk, H.U.H., Bangalore, S., Wang, Z., Johnson, K.W., Pinotti, R., Zhang, H., Kaplin, S., Narasimhan, B., Kitai, T., et al.: Machine learning prediction in cardiovascular diseases: a meta-analysis. Scientific reports 10(1), 16057 (2020)

[8] Topol, E.J.: High-performance medicine: the convergence of human and artificial intelligence. Nature medicine 25(1), 44–56 (2019)

[9] Kumari, A., Punn, N.S., Sonbhadra, S.K., Agarwal, S.: Impact of the composition of feature extraction and class sampling in medicare fraud detection. In: International Conference on Neural Information Processing, pp. 639–658 (2022). Springer

[10] Nagabhushan, P., Sonbhadra, S.K., Punn, N.S., Agarwal, S.: Towards machine learning to machine wisdom: a potential quest. In: International Conference on Big Data Analytics, pp. 261–275 (2021). Springer

[11] Carvalho, D., Cruz, R.: Big data and machine learning in health. European Journal of Public Health 30(Supplement 2), 040–030 (2020)

[12] Opoku-Acheampong, A.A., Rosenkranz, R.R., Adhikari, K., Muturi, N., Logan, C., Kidd, T.: Tools for assessing cardiovascular disease risk factors in underserved young adult populations: a systematic review. International Journal of Environmental Research and Public Health 18(24), 13305 (2021)

[13] Wong, Y.-K., Tse, H.-F.: Circulating biomarkers for cardiovascular disease risk prediction in patients with cardiovascular disease. Frontiers in Cardiovascular Medicine 8, 713191 (2021)

[14] Sagi, O., Rokach, L.: Ensemble learning: A survey. Wiley Interdisciplinary Reviews: Data Mining and Knowledge Discovery 8(4), 1249 (2018)

[15] Dimopoulos, A.C., Nikolaidou, M., Caballero, F.F., Engchuan, W., Sanchez-Niubo, A., Arndt, H., Ayuso-Mateos, J.L., Haro, J.M., Chatterji, S., Georgousopoulou, E.N., et al.: Machine learning methodologies versus cardiovascular risk scores, in predicting disease risk. BMC medical research methodology 18, 1–11 (2018)

[16] Hassan, C.A.u., Iqbal, J., Irfan, R., Hussain, S., Algarni, A.D., Bukhari, S.S.H., Alturki, N., Ullah, S.S.: Effectively predicting the presence of coronary heart disease using machine learning classifiers. Sensors 22(19), 7227 (2022)

[17] Pajila, P.B., Sheena, B.G., Gayathri, A., Aswini, J., Nalini, M., et al.: A comprehensive survey on naive bayes algorithm: Advantages, limitations and applications. In: 2023 4th International Conference on Smart Electronics and Communication (ICOSEC), pp. 1228–1234 (2023). IEEE

[18] Khateeb, N., Usman, M.: Efficient heart disease prediction system using k-nearest neighbor classification technique. In: Proceedings of the International Conference on Big Data and Internet of Thing, pp. 21–26 (2017)

[19] Wisaeng, K.: Predict the diagnosis of heart disease using feature selection and k-nearest neighbor algorithm. Applied Mathematical Sciences 8(83), 4103–4113 (2014)

[20] Parmar, A., Katariya, R., Patel, V.: A review on random forest: An ensemble classifier. In: International Conference on Intelligent Data Communication Technologies and Internet of Things (ICICI) 2018, pp. 758–763 (2019). Springer

[21] Mohan, S., Thirumalai, C., Srivastava, G.: Effective heart disease prediction using hybrid machine learning techniques. IEEE access 7, 81542–81554 (2019)

[22] Yang, L., Wu, H., Jin, X., Zheng, P., Hu, S., Xu, X., Yu, W., Yan, J.: Study of cardiovascular disease prediction model based on random forest in eastern china. Scientific reports 10(1), 5245 (2020)

[23] Sudhanshu Punn, N.S., Sonbhadra, S.K., Agarwal, S.: Recommending best course of treatment based on similarities of prognostic markers. In: Neural Information Processing: 28th International Conference, ICONIP 2021, Sanur, Bali, Indonesia, December 8–12, 2021, Proceedings, Part II 28, pp. 393–404 (2021). Springer

[24] Vettoruzzo, A., Bouguelia, M.-R., Vanschoren, J., Rognvaldsson, T., Santosh, K.: Advances and challenges in meta-learning: A technical review. IEEE Transactions on Pattern Analysis and Machine Intelligence (2024)

[25] Vilalta, R., Drissi, Y.: A perspective view and survey of meta-learning. Artificial intelligence review 18, 77–95 (2002)

[26] Lemke, C., Budka, M., Gabrys, B.: Metalearning: a survey of trends and technologies. Artificial intelligence review 44, 117–130 (2015)

[27] Weng, S.F., Reps, J., Kai, J., Garibaldi, J.M., Qureshi, N.: Can machine-learning improve cardiovascular risk prediction using routine clinical data? PloS one 12(4), 0174944 (2017)

[28] Ghosh, P., Azam, S., Jonkman, M., Karim, A., Shamrat, F.J.M., Ignatious, E., Shultana, S., Beeravolu, A.R., De Boer, F.: Efficient prediction of cardiovascular disease using machine learning algorithms with relief and lasso feature selection techniques. IEEE Access 9, 19304–19326 (2021)

[29] Sun, Q., Liu, Y., Chen, Z., Chua, T.-S., Schiele, B.: Meta-transfer learning through hard tasks. IEEE Transactions on Pattern Analysis and Machine Intelligence 44(3), 1443–1456 (2020)

[30] Gao, M., Jiang, H., Zhu, L., Jiang, Z., Geng, M., Ren, Q., Lu, Y.: Discriminative ensemble meta-learning with co-regularization for rare fundus diseases diagnosis. Medical Image Analysis 89, 102884 (2023)

[31] Tsao, C.W., Aday, A.W., Almarzooq, Z.I., Anderson, C.A., Arora, P., Avery, C.L., Baker-Smith, C.M., Beaton, A.Z., Boehme, A.K., Buxton, A.E., et al.: Heart disease and stroke statistics—2023 update: a report from the american heart association. Circulation 147(8), 93–621 (2023)

[32] Liu, W., Laranjo, L., Klimis, H., Chiang, J., Yue, J., Marschner, S., Quiroz, J.C., Jorm, L., Chow, C.K.: Machine-learning versus traditional approaches for atherosclerotic cardiovascular risk prognostication in primary prevention cohorts: a systematic review and meta-analysis. European Heart Journal-Quality of Care and Clinical Outcomes 9(4), 310–322 (2023)

[33] Marbaniang, I.A., Choudhury, N.A., Moulik, S.: Cardiovascular disease (cvd) prediction using machine learning algorithms. In: 2020 IEEE 17th India Council International Conference (INDICON), pp. 1–6 (2020). IEEE

[34] Kaggle: Cardiovascular Disease Dataset. [Online; accessed September, 2023] (2019). https://www.kaggle.com/datasets/sulianova/cardiovascular-disease-dataset

